# Pre-pandemic mental and physical health as predictors of COVID-19 vaccine hesitancy: evidence from a UK-wide cohort study

**DOI:** 10.1101/2021.04.27.21256185

**Authors:** G. David Batty, Ian J. Deary, Drew Altschul

## Abstract

**Importance:** Although several predictors of COVID-19 vaccine hesitancy have been identified, the role of physical health has not been well-examined, and the association with mental health is unknown.

**Objective:** To examine the association of pre-pandemic mental health, physical health, and shielding with vaccine hesitancy after the announcement of the successful testing of the Oxford University/AstraZeneca vaccine.

**Design, Setting, and Participants:** We used individual-level data from a pandemic-focused investigation (COVID Survey), a prospective cohort study nested within the UK Understanding Society (Main Survey) project. In the week immediately following the announcement of successful testing of the first efficacious inoculation (November/December 2020), data on vaccine intentionality were collected in 12,035 individuals aged 16-95 years. Pre-pandemic, study members had responded to enquiries about diagnoses of mental and physical health, completed the 12-item General Health Questionnaire for symptoms of psychological distress (anxiety and depression), and indicated whether they or someone in their household was shielding.

**Main outcome measures:** Self-reported intention to take up a vaccination for COVID-19. To summarise our results, we computed odds ratios with accompanying 95% confidence intervals for indices of health and shielding adjusted for selected covariates.

**Results:** In an analytical sample of 11,955 people (6741 women), 15.4% indicated that they were vaccine hesitant. Relative to their disease-free counterparts, shielding was associated with a 24% lower risk of being hesitant (odds ratio; 95% confidence interval: 0.76; 0.59, 0.96), after adjustment for a range of covariates which included age, education, and ethnicity. Corresponding results for cardiometabolic disease were 22% (0.78; 0.64, 0.95), and for respiratory disease were 26% (0.74; 0.59, 0.93). Having a pre-pandemic diagnosis of anxiety or depression, or a high score on the distress symptom scale, were all unrelated to the willingness to take up a vaccine.

**Conclusions and relevance:** People who have been prioritised for COVID-19 vaccination owing to a physical condition are more likely to take it up. These effects were not apparent for indices of mental health.

## Introduction

Whereas it was established early in the COVID-19 pandemic that people with chronic physical illness experienced higher rates of hospitalisation for and death from the disease,^1-4^ more recent evidence suggests that the same may also be the case for people with a mental health problems and those with a higher prevalence of psychological distress symptoms (anxiety and depression).^4-7^ In selected countries, vaccination against COVID-19, central to attaining herd immunity,^8^ has therefore been prioritised for people with physical disorders – alongside the request that they shield – whereas population segmentation of people with mental health problems has been much less universal.^9^

There are several reasons to anticipate greater vaccine hesitancy in people with mental health problems, including those with high scores on distress symptom scale, and this may compound COVID-19 rates in this at-risk group. First, individuals with psychiatric morbidity and symptoms of distress tend to have a lower prevalence of health-protecting behaviours. Relative to their unaffected counterparts, for instance, they are more likely to smoke, take less exercise, have an imprudent diet, and be obese.^10-12^ Third, people with mental health issues also appear to be less likely to take up the offer of health screening,^13^ although this is not a universal observation.^14^ Lastly, of most relevance, in a study of influenza inoculation, users of an outpatient psychiatry clinic had markedly lower take up than the general population.^15^

Collectively, these observations provide a *prima facie* case that people with common symptoms of mental distress may be somewhat more hesitant when offered a vaccination against COVID-19, and in the absence of any empirical data, there have been recent calls to test this relationship.^16^ Accordingly, in a large, general-population based UK sample we examined the relationships of mental health diagnosis and symptoms of mental distress with vaccine hesitancy. Alongside these results, we present the association between somatic illness and vaccine hesitancy; in the few relevant studies, having somatic illness has been significantly associated with lower levels of hesitancy.^17,18^ Importantly, data collection on vaccine intention took place following the announcement of successful testing of the Oxford University/AstraZeneca vaccine, which was widely and prominently publicised. Therefore, the present survey concerning vaccination hesitancy was taken at a time when the future offer of a vaccination was no longer merely hypothetical.

## Methods

Understanding Society, also known as the UK Household Longitudinal Study, is a nationally-representative, on-going, open, cohort study (hereinafter, the ‘Main Survey’). Based on a clustered-stratified probability sample of households, participants have been interviewed annually since 2009.^19^ Households who had participated in at least one of the two most recent waves of data collection (wave 8, 2016-18; wave 9, 2017-19) comprised the target sample for a pandemic-focused study initiated in April 2020 (hereinafter, the ‘COVID Survey’).^20,21^ The derivation of the present analytical sample from the Main and COVID Surveys, including the wave for specific data collection, is given in figure 1. The University of Essex Ethics Committee gave approval for the COVID-orientated surveys (ETH1920-1271); no further ethical permissions were required for the present analyses of anonymised data.

**Figure 1.**
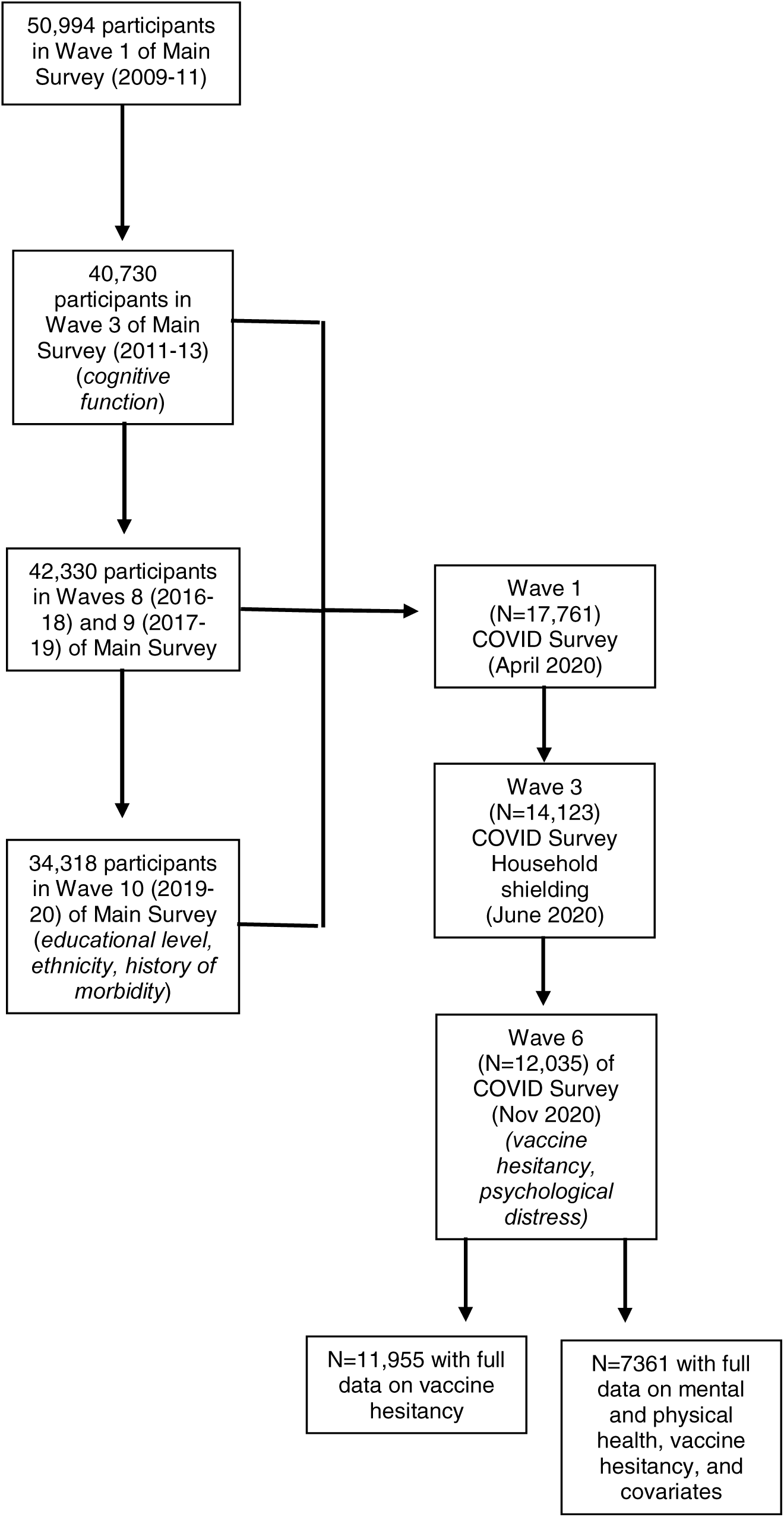
Flow of cohort members into the analytical sample: Main Survey and COVID Survey in Understanding Society

The COVID Surveys took place monthly/bimonthly between April (wave 1) and November 2020 (wave 6), with questions on vaccine intention first administered in the latest tranche of data collection when study members were aged 16-95 years (mean 53).^21^ Data collection in wave 6 (starting 24th November) commenced the day immediately following the announcement of the efficacy of the Oxford University/AstraZeneca vaccine.^22^ Data collection continued for one week, and obtained information from a total of 12,035 individuals of 19,294 invitations issued (response proportion 62%).^21^

### Assessment of mental and physical morbidity

Study members indicated if a physician or other health professional had ever informed them that they had a psychiatric problem, which included anxiety, depression, psychosis or schizophrenia, bipolar disorder or manic depression, an eating disorder, post-traumatic stress disorder, or any other mental illness (wave 10, 2019-20; Main Survey). With a low prevalence hesitancy for selected conditions, we aggregated the latter five mental health groups. Self-reports of a physician diagnosis of mental illness, in particular depression, shows reasonable agreement with structured clinical interviews.^23^

Psychological distress (wave 6, November 2020; COVID Survey) was ascertained using administration of the 12-item version of the General Health Questionnaire. Validated against standardised psychiatric interviews,^24,25^ this is a widely-used measure of psychological distress in population-based studies. Consistent with published analyses,^12,26,27^ we used the following classifications: asymptomatic (score 0), sub-clinically symptomatic (score 1-3), symptomatic (score 4-6), and highly symptomatic (score 7-12).

A history of physical morbidity was also captured (wave 10, 2019-20; Main Survey) and based on self-report of physician diagnosis for: a cardiometabolic condition (congestive heart failure, coronary heart disease, angina, heart attack or infarction, stroke, diabetes, and/or hypertension); respiratory disease (respiratory disease comprised bronchitis, emphysema, chronic obstructive pulmonary disease, and/or asthma); or cancer of any type. In other studies, these data reveal moderate to high agreement with clinical records.^28^

Lastly, based on their physical medical history, people judged as extremely clinically vulnerable to COVID-19 were contacted by the UK National Health Service or their general practitioner during the early stages of the pandemic and recommended to stay at home. Conditions that met the criteria for shielding included selected cancers, severe respiratory disorders such as cystic fibrosis, severe asthma, organ transplant recipients, and people with a disability such as Down’s syndrome.^29^ Study members were asked about the shielding status for themselves or a household member (waves 1-5, April to July 2020; COVID Surveys; denoted by yes/no).

### Assessment of covariates

Covariates were self-reported and included age; sex (both wave 10, 2019-20; Main Survey); ethnicity (wave 10, Main Survey; denoted as white or non-white); and highest education level (wave 10, Main Survey; categorised as degree & other higher degree, A’ level or equivalent [Advanced Placement in the USA], GCSE or equivalent [Grade 10 in the USA], other qualification, and none). In the third wave of data collection in the Main Survey (2011-2013), six cognitive function tests were administered: immediate word recall and delayed word recall tasks; semantic verbal fluency; cognitive impairment; numerical reasoning skills; and fluid reasoning.^30^ Representing a range of cognitive skills, these tests have been repeatedly deployed in large-scale, population-based studies.^31-35^ Using scores from the six tests, we generated a single general cognitive function variable (*g*) for use in the present analyses.^36^

### Assessment of vaccine hesitancy

At wave 6 (November 2020) in the COVID Survey, study members were asked: “Imagine that a vaccine against COVID-19 was available for anyone who wanted it. How likely or unlikely would you be to take the vaccine?”. Possible responses were “Very likely”, “Likely”, “Unlikely” and “Very unlikely”. The latter two categories were combined to denote vaccine hesitancy.

### Statistical analyses

To summarise the relation between mental morbidity, physical morbidity, and vaccine hesitancy, we used logistic regression to compute odds ratios with accompanying 95% confidence intervals. The most basic analyses were adjusted for age, sex, and ethnicity. Retaining these covariates, we then explored the impact of controlling separately and collectively for education, shielding status, and cognitive function; in analyses in which mental health was the exposure of interest, we physical illness, and vice versa.

## Results

In table 1 we show study member characteristics according to vaccine intention in unadjusted analyses. In a sample of 11,955 individuals (6741 women) who responded in full to the enquiry regarding COVID-19 vaccine intentionality, 15.4% indicated that they were hesitant. Relative to the group who indicated a willingness to have the vaccine, those who were hesitant were more likely to be younger, female, from an ethnic minority background, be less well educated, and have a lower general cognitive function score. The hesitant were also less likely to have an existing somatic morbidity, as indexed by cardiometabolic disease and cancer. Related, there was also a lower prevalence of shielding in the hesitant category (correlation between any physical morbidity and shielding in the present study: ρ=0.12, p<0.0001, N=10916). There was, however, little evidence of a difference in prevalence of specific mental health diagnoses across the hesitant groups; only ‘other’ mental health conditions was more common in study members expressing hesitancy, but the absolute difference was marginal with statistical significance generated from the large numbers. People who declared themselves reticent in taking the vaccine when offered had slightly higher levels of psychological distress symptoms.

**Table 1.**
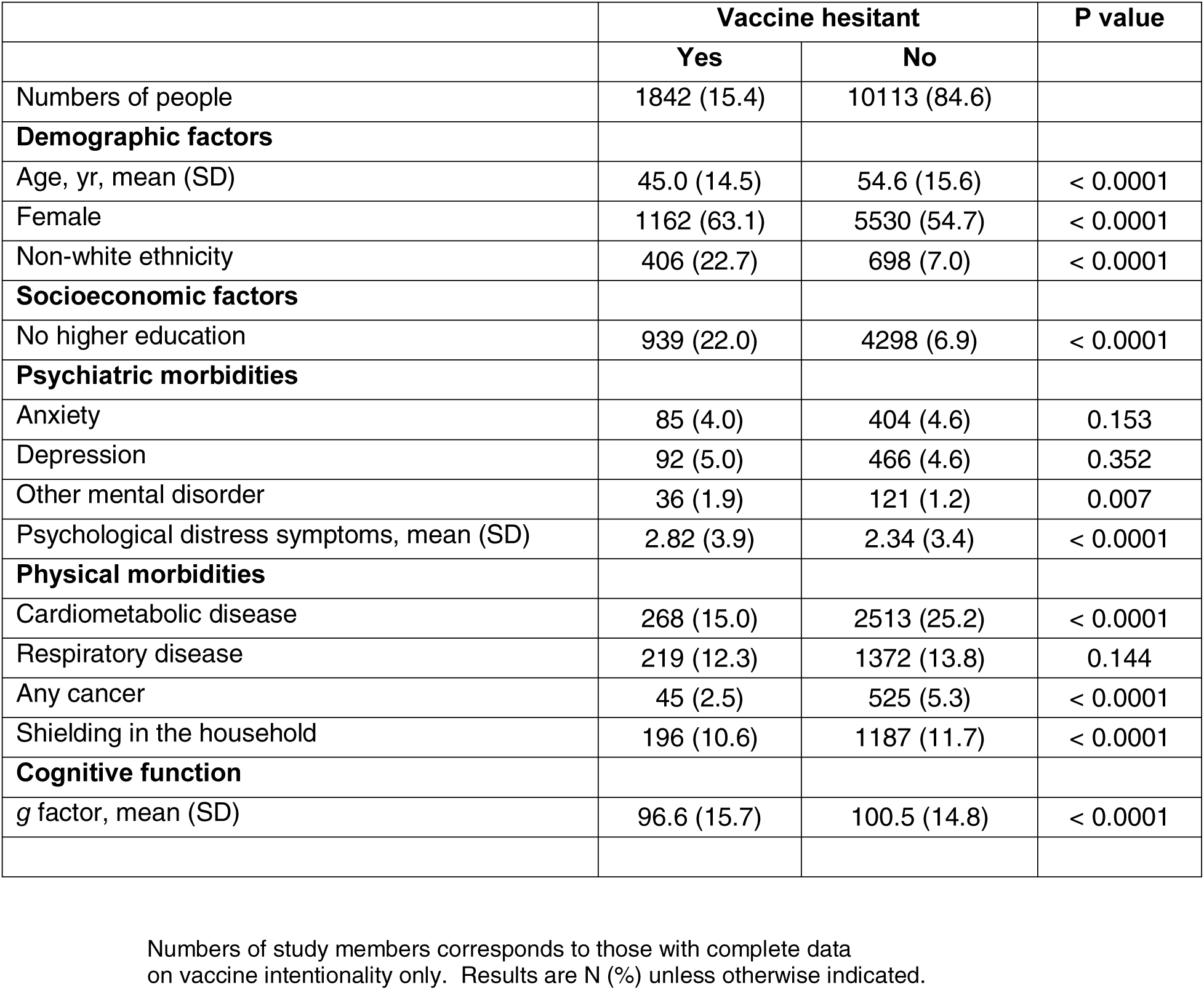
Study member characteristics according to COVID-19 vaccine hesitancy in Understanding Society

|  | Vaccine hesitant |  | P value |
| --- | --- | --- | --- |
|  | Yes | No |  |
| Numbers of people | 1842 (15.4) | 10113 (84.6) |  |
| **Demographic factors** |  |  |  |
| Age, yr, mean (SD) | 45.0 (14.5) | 54.6 (15.6) | < 0.0001 |
| Female | 1162 (63.1) | 5530 (54.7) | < 0.0001 |
| Non-white ethnicity | 406 (22.7) | 698 (7.0) | < 0.0001 |
| **Socioeconomic factors** |  |  |  |
| No higher education | 939 (22.0) | 4298 (6.9) | < 0.0001 |
| **Psychiatric morbidities** |  |  |  |
| Anxiety | 85 (4.0) | 404 (4.6) | 0.153 |
| Depression | 92 (5.0) | 466 (4.6) | 0.352 |
| Other mental disorder | 36 (1.9) | 121 (1.2) | 0.007 |
| Psychological distress symptoms, mean (SD) | 2.82 (3.9) | 2.34 (3.4) | < 0.0001 |
| **Physical morbidities** |  |  |  |
| Cardiometabolic disease | 268 (15.0) | 2513 (25.2) | < 0.0001 |
| Respiratory disease | 219 (12.3) | 1372 (13.8) | 0.144 |
| Any cancer | 45 (2.5) | 525 (5.3) | < 0.0001 |
| Shielding in the household | 196 (10.6) | 1187 (11.7) | < 0.0001 |
| **Cognitive function** |  |  |  |
| g factor, mean (SD) | 96.6 (15.7) | 100.5 (14.8) | < 0.0001 |

Numbers of study members corresponds to those with complete data on vaccine intentionality only. Results are N (%) unless otherwise indicated.

In table 2 we used multiple regression analyses to explore the association between an existing diagnosis of a morbidity as a predictor of vaccine hesitancy. Relative to people without a physical condition, those with a diagnosis of cardiometabolic disease (odds ratio; 95% confidence interval: 0.82; 0.67, 0.99) or respiratory disease (0.71; 0.57, 0.88) were less like to have reported that they would decline an offer of vaccination, after adjustment for age, sex, and ethnicity. The associations between cancer and shielding and vaccine hesitancy were not statistically significant at conventional levels. Adjusting for a range of covariates (table 2 and figure 2) had little impact on these relationships; an exception was the regression coefficient for shielding becoming statistically significant at conventional levels (0.76; 0.59, 0.96) such that people who were shielding were less vaccine-hesitant. The general lack of impact of control for individual covariates is shown in table a1 (appendix).

**Table 2.** Odds ratios (95% confidence intervals) for the relation of mental and physical health with later COVID-19 vaccine hesitancy in Understanding Society (N=7361)

|  | Number hesitant<br>/ Total at risk | Age, sex, & ethnicity | All covariates |
| --- | --- | --- | --- |
| **Psychiatric morbidity** |  |  |  |
| Anxiety | 50/324 | 1.00 (0.72, 1.36) | 1.11 (0.79, 1.52) |
| Depression | 54/368 | 0.99 (0.72, 1.33) | 1.12 (0.81, 1.53) |
| Other mental health condition(s) | 20/111 | 1.08 (0.64, 1.75) | 1.21 (0.71, 1.97) |
| Any mental health condition | 71/491 | 0.99 (0.75, 1.29) | 1.14 (0.86, 1.49) |
| **Psychological distress** |  |  |  |
| Asymptomatic (score 0) | 443/3339 | 1.0 (ref) | 1.0 (ref) |
| Subclinically symptomatic (1-3) | 247/2256 | 0.77 (0.64, 0.91) | 0.81 (0.63, 0.98) |
| Symptomatic (4-6) | 90/750 | 0.77 (0.59, 0.98) | 0.82 (0.56, 1.07) |
| Highly symptomatic (7-12) | 173/1016 | 1.05 (0.85, 1.28) | 1.12 (0.92, 1.33) |
| P for quadratic association |  | < 0.0001 | 0.003 |
| P for linear trend |  | 0.251 | 0.075 |
| Per SD (3.5 points) decrease | 953/7361 | 0.93 (0.81, 1.06) | 0.88 (0.75, 1.02) |
| **Physical morbidity** |  |  |  |
| Cardiometabolic disease | 147/1905 | 0.82 (0.67, 0.99) | 0.78 (0.64, 0.95) |
| Respiratory disease | 107/1034 | 0.71 (0.57, 0.88) | 0.74 (0.59, 0.93) |
| Any cancer | 29/389 | 0.87 (0.58, 1.28) | 0.95 (0.62, 1.39) |
| Any physical health condition | 225/2389 | 0.72 (0.61, 0.85) | 0.72 (0.60, 0.85) |
| Shielding in household | 88/889 | 0.81 (0.63, 1.03) | 0.76 (0.59, 0.96) |

All covariates are: age, sex, ethnicity, education, shielding status, and cognitive function. Effect estimates for physical morbidity and psychiatric morbidity were mutually-adjusted. Psychological distress symptoms were ascertained from the General Health Questionnaire.

**Figure 2.**
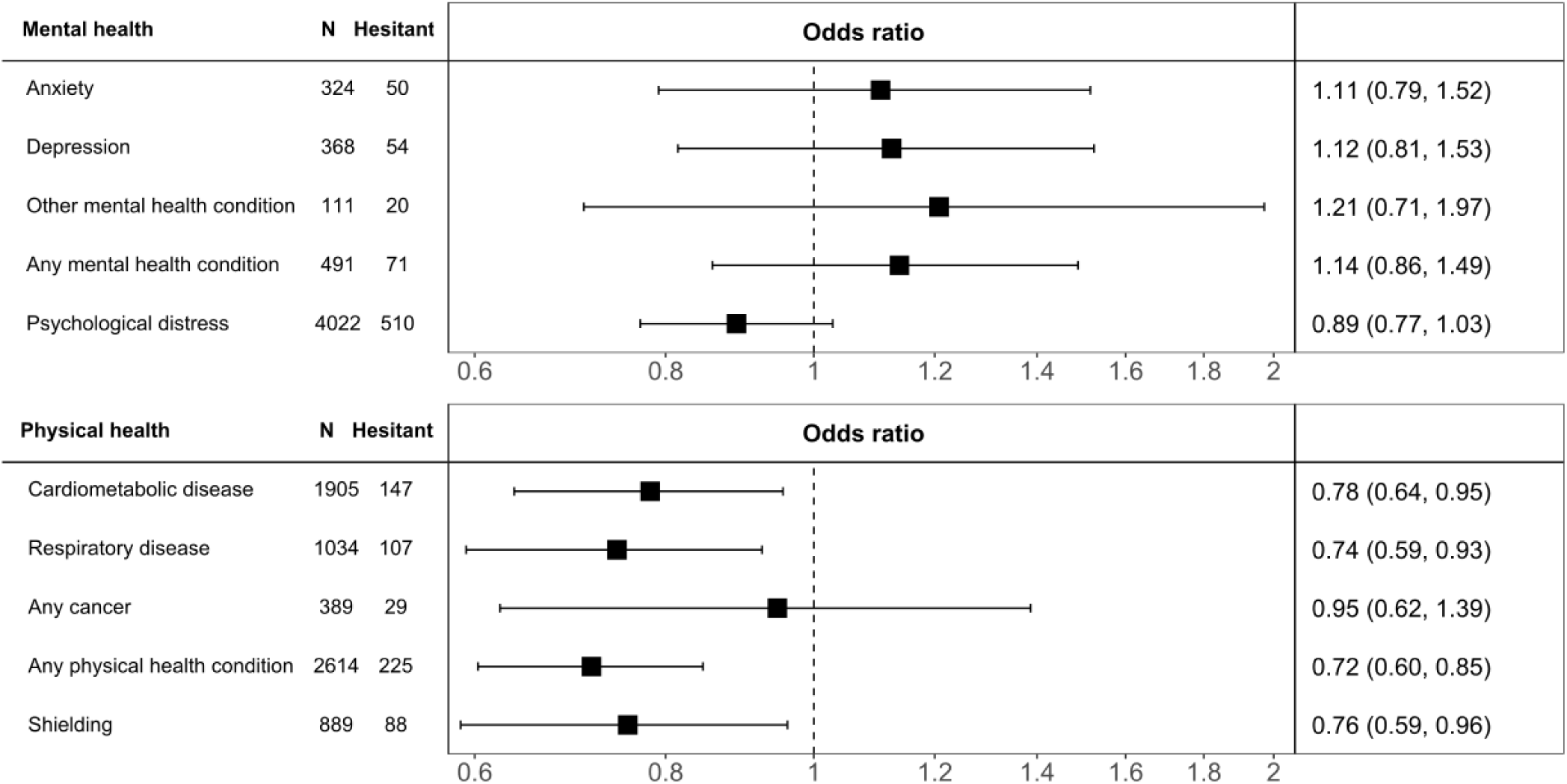
Odds ratios (95% confidence intervals) for the relation of mental and physical health with later COVID-19 vaccine hesitancy in Understanding Society (N=7361) Numbers of study members in this sample corresponds to those with complete data on all variables in the analyses. Effect estimates for physical morbidity and psychiatric morbidity were mutually-adjusted. For each morbidity, the referent group is those study members without the condition.

In analyses in which mental illness diagnosis was the exposure of interest, none of the individual psychiatric conditions were related to vaccine hesitancy (table 2). Using the standard four category schema for symptoms of psychological distress, however, there was some suggestion of a ‘U’-shaped effect, such that people who had either low or high scores on the distress scale were marginally more likely to be vaccine hesitant, and those with moderate symptoms had the lowest likelihood (p-value for quadratic relationship after multiple adjustment: 0.003). We further explored this association by using raw scores from the psychological distress scale (range 0-12). Based on this disaggregation, there was, however, no support for any relationship, linear or quadratic, between psychological distress and vaccine hesitancy (figure 3).

**Figure 3.**
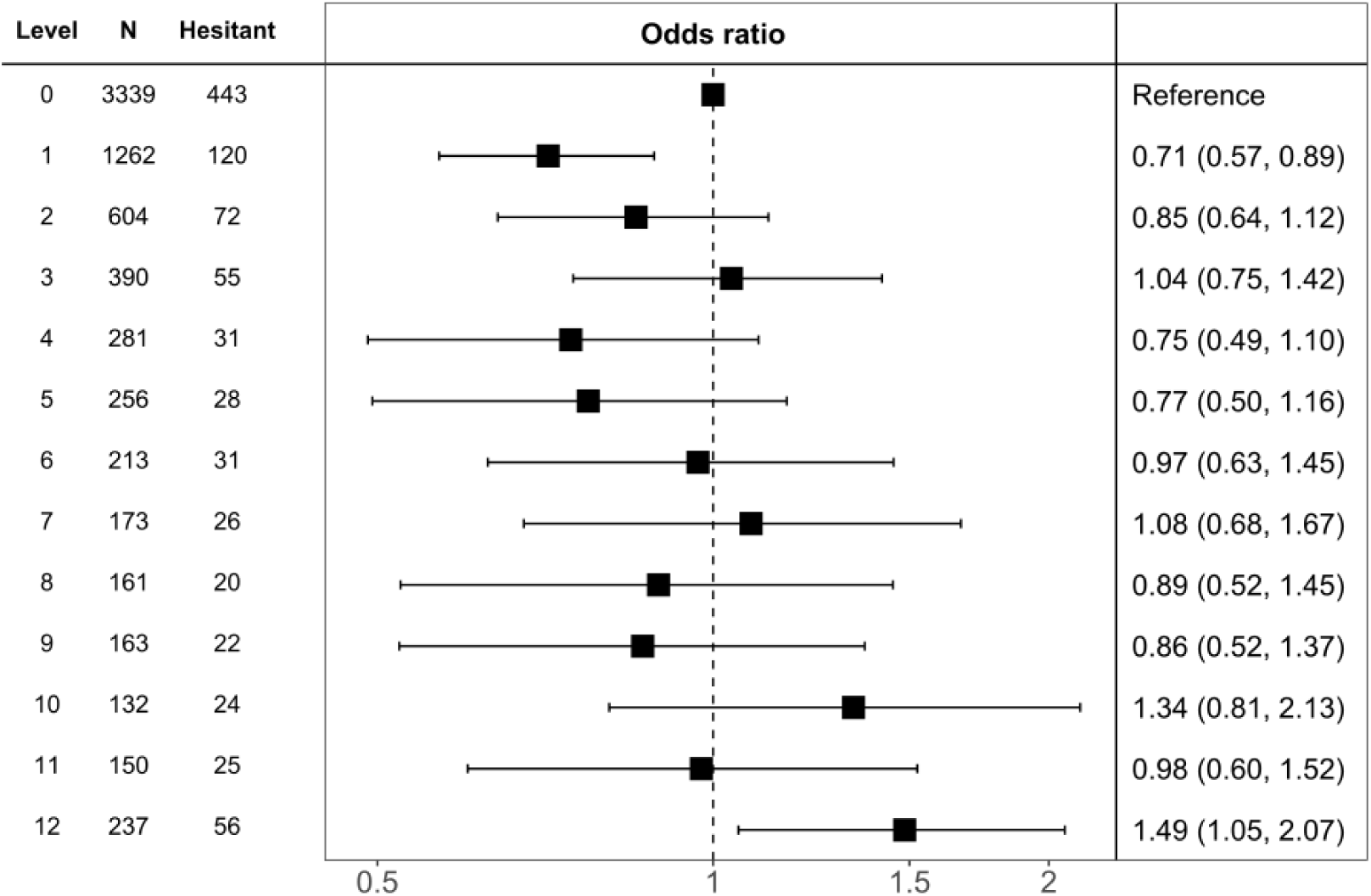
Multiply-adjusted odds ratios (95% confidence intervals) for the relation of psychological distress with later COVID-19 vaccine hesitancy in Understanding Society (N=7361) All covariates are: age, sex, ethnicity, education, somatic comorbidity, shielding, and cognitive function.

## Discussion

Our main finding was that, in data collected in the United Kingdom immediately following the announcement of the successful evaluation of the Oxford University/AstraZeneca vaccine, selected physical but not psychiatric morbidities were related to a lower likelihood of vaccine hesitancy. The results for mental health run counter to our expectations, given that people with such morbidities are, as described, less likely to engage in health-protecting behaviours such as healthy lifestyle habits^12^ and screening for somatic disorder.^13^

### Comparison with existing studies

The notion that people with a long-standing physical condition are less likely to be vaccine-hesitant has been reported in other studies.^17,18^ That we also recapitulated known associations with hesitancy such as being female,^37-39^ being younger,^37,39^ and from an ethnic minority group,^21,39,40^ gives us some confidence in our novel results for mental health. To the best of our knowledge, there has been one prior examination of the relationship between mental health and vaccine hestinacy.^41^ Comprising two small cross-sectional studies where data collection took place prior to the announcement of the successful testing of the first efficacious vaccination, study members were administered a very brief and unvalidated enquiry as to whether they had experience of mental health problems. In that study, there was no clear evidence of a link.^41^ Studies using data based on other vaccination programmes offer limited insights. For instance, in a cross-sectional study of patients with schizophrenia which took place during the 2009 H1N1 influenza pandemic in Australia, three-quarters indicated that they were willing to be vaccinated;^42^ however, in keeping with similar studies,^43^ the absence of a general population comparison group renders interpretation problematic. In a small cohort of socioeconomically disadvantaged mothers, those with mental health problems were seemingly less likely to have children with up-to-date vaccine coverage, although the association was weak and the study underpowered.^44^

### Study strengths and weaknesses

Whereas the present study has its strengths, including its size, national representativeness, and timing, there are also some weaknesses. First, we used vaccine intentionality as an indicator vaccine uptake but the correlation between the two is imperfect. In a small-scale longitudinal study conducted during the period of the 2009 H1N1 pandemic in Hong Kong, less than 10% of people who expressed a commitment to being inoculated reported that they had received a vaccination two months later.^45^ Elsewhere, in a US adult population at high risk of seasonal influenza, around half of those intending to be vaccinated had received the inoculation within the following 5 months.^46^ Second, there was inevitably some loss to follow-up (figure 1). Whereas this attrition might have impacted upon the estimation of the prevalence vaccine hesitancy, which is likely to be lower in our select sample relative to the general population,^47^ it is unlikely to have influenced our estimation of its relationship with mental and physical health. Thus, in other contexts, we have shown that highly-selected cohorts reveal very similar risk factor–outcome associations to those seen in studies with conventionally high response.^48^ In conclusion, we found that some somatic conditions but not mental health problems were related to a lower likelihood of being vaccine hesitant against COVID-19.

## Data Availability

Researchers who would like to use Understanding Society data need to register with the UK Data Service (https://www.ukdataservice.ac.uk/) before application.

## Appendix

*Batty GD, Deary IJ, Altschul D. Pre-pandemic mental and physical Health as predictors of COVID-19 vaccine hesitancy: evidence from a UK cohort study*

**Table a1.** Odds ratios (95% confidence intervals) for the relation of mental and physical health with later COVID-19 vaccine hesitancy in Understanding Society – with models featuring individual covariates (N=7361)

|  | Number<br>hesitant /<br>Total at<br>risk | Age, sex, &<br>ethnicity | Age, sex,<br>ethnicity, &<br>comorbidity | Age, sex,<br>ethnicity, &<br>shielding | Age, sex,<br>ethnicity, &<br>education | Age, sex,<br>ethnicity, and<br>cognition | All covariates |
| --- | --- | --- | --- | --- | --- | --- | --- |
| **Psychiatric morbidity** |  |  |  |  |  |  |  |
| Anxiety | 50/324 | 1.00 (0.72, 1.36) | 1.06 (0.76, 1.44) | 1.00 (0.72, 1.36) | 1.04 (0.74, 1.42) | 1.02 (0.73, 1.39) | 1.11 (0.79, 1.52) |
| Depression | 54/368 | 0.99 (0.72, 1.33) | 1.05 (0.77, 1.43) | 0.99 (0.72, 1.34) | 1.02 (0.74, 1.38) | 1.03 (0.75, 1.39) | 1.12 (0.81, 1.53) |
| Other mental health condition(s) | 20/111 | 1.08 (0.64, 1.75) | 1.17 (0.69, 1.89) | 1.08 (0.64, 1.75) | 1.06 (0.62, 1.73) | 1.15 (0.68, 1.87) | 1.21 (0.71, 1.97) |
| Any mental health condition | 71/491 | 0.99 (0.75, 1.29) | 1.05 (0.79, 1.37) | 0.99 (0.75, 1.29) | 1.04 (0.78, 1.35) | 1.05 (0.79, 1.37) | 1.14 (0.86, 1.49) |
| **Psychological distress** |  |  |  |  |  |  |  |
| Asymptomatic (score 0) | 443/3339 | 1.0 (ref) | 1.0 | 1.0 | 1.0 | 1.0 | 1.0 |
| Subclinically symptomatic (1-3) | 247/2256 | 0.77 (0.64, 0.91) | 0.77 (0.68, 0.96) | 0.77 (0.60, 0.94) | 0.79 (0.62, 0.97) | 0.79 (0.62, 0.96) | 0.81 (0.63, 0.98) |
| Symptomatic (4-6) | 90/750 | 0.77 (0.59, 0.98) | 0.78 (0.63, 1.05) | 0.77 (0.52, 1.02) | 0.78 (0.53, 1.04) | 0.78 (0.53, 1.04) | 0.82 (0.56, 1.07) |
| Highly symptomatic (7-12) | 173/1016 | 1.05 (0.85, 1.28) | 1.08 (0.91, 1.38) | 1.06 (0.86, 1.26) | 1.07 (0.87, 1.27) | 1.07 (0.86, 1.27) | 1.12 (0.92, 1.33) |
| P for quadratic |  | < 0.0001 | 0.017 | 0.028 | 0.034 | 0.036 | 0.003 |
| P for linear trend |  | 0.251 | 0.099 | 0.148 | 0.147 | 0.145 | 0.075 |
| Per SD (3.5 points) decrease | 953/7361 | 0.93 (0.81, 1.06) | 0.91 (0.79, 1.04) | 0.92 (0.80, 1.05) | 0.92 (0.80, 1.05) | 0.92 (0.80, 1.05) | 0.88 (0.75, 1.02) |
| **Physical morbidity** |  |  |  |  |  |  |  |
| Cardiometabolic disease | 147/1905 | 0.82 (0.67, 1.00) | 0.82 (0.67, 1.00) | 0.83 (0.68, 1.01) | 0.80 (0.66, 0.98) | 0.78 (0.64, 0.95) | 0.78 (0.64, 0.95) |
| Respiratory disease | 107/1034 | 0.71 (0.57, 0.88) | 0.71 (0.57, 0.88) | 0.72 (0.58, 0.90) | 0.73 (0.58, 0.90) | 0.72 (0.57, 0.89) | 0.74 (0.59, 0.93) |
| Any cancer | 29/389 | 0.87 (0.58, 1.28) | 0.87 (0.58, 1.28) | 0.89 (0.59, 1.30) | 0.90 (0.59, 1.32) | 0.92 (0.61, 1.34) | 0.95 (0.62, 1.39) |
| Any physical health condition | 225/2389 | 0.72 (0.61, 0.85) | 0.72 (0.61, 0.85) | 0.73 (0.61, 0.86) | 0.72 (0.61, 0.85) | 0.71 (0.60, 0.83) | 0.72 (0.60, 0.85) |
| Shielding in household | 196/1383 | 0.81 (0.63, 1.03) | 0.81 (0.63, 1.03) |  | 0.78 (0.61, 1.00) | 0.76 (0.59, 0.97) | 0.76 (0.59, 0.96) |

All covariates are: age, sex, ethnicity, education, shielding, and cognitive function. Effect estimates for physical morbidity and psychiatric morbidity were mutually-adjusted

## Notes

Funding: GDB is supported by the UK Medical Research Council (MR/P023444/1) and the US National Institute on Aging (1R56AG052519-01; 1R01AG052519-01A1); and IJD by the UK Medical Research Council (MR/R024065/1), UK Economic and Social Research Council (ES/S015604/1), and the US National Institute on Aging (1R01AG054628-01A1). These funders, who provided no direct financial or material support for the work, had no role in study design, data collection, data analysis, data interpretation, or report preparation.

### Competing Interest Statement

The authors have declared no competing interest.

### Funding Statement

GDB is supported by the UK Medical Research Council (MR/P023444/1) and the US National Institute on Aging (1R56AG052519-01; 1R01AG052519-01A1); and IJD by the UK Medical Research Council (MR/R024065/1), UK Economic and Social Research Council (ES/S015604/1), and the US National Institute on Aging (1R01AG054628-01A1). These funders, who provided no direct financial or material support for the work, had no role in study design, data collection, data analysis, data interpretation, or report preparation.

### Author Declarations

The University of Essex Ethics Committee gave approval for the COVID-orientated surveys (ETH1920-1271); no further ethical permissions were required for the present analyses of anonymised data.

